# Mortality Predictions for Men and Women Diagnosed with Anal Canal Squamous Cell Carcinoma in the United States

**DOI:** 10.1101/2023.02.05.23285488

**Authors:** Roungu Ahmmad, Pegah Hosseini-Carroll, Sanwar Hossain, Raina Muthukumaru

## Abstract

**Objective:** Studies have concluded that treatments for patients with anal cancer vary based on their age, sex, and race. We examined gender-specific differences in mortality and overall survival for patients with squamous cell carcinoma (SCC) of the anal canal (AC), based on differences in treatment, stage and demographic factors.

**Method:** SEER’s retrospective cohort dataset was used to review all patients diagnosed with SCC of the AC between 2000 and 2018. We analyzed survival by sub-distributed Cox models and estimated overall survival probabilities through nomograms based on cancer treatments, tumor characteristics, and demographic data

**Results:** In the study, 24,892 patients with SCC in the AC had a median survival time of 43 months (IQR: 16-97). However, males (median 38.0 months, IQR: 13.0-92.0) had significantly shorter median survival times than females (median 46.0 months, IQR: 17-100). There were 12% of patients in metastatic stages, with females (70.3%) receiving more treatments than males (29.7%). The survival rate of patients with SCC of the AC who underwent radiation treatment was significantly higher (HR 0.72, 95% CI: 0.65-0.80, p<0.001). Moreover, compared to those without surgery on the primary site, those undergoing surgery on the primary site had a 49% lower mortality rate (HR 0.51; 95% CI: 0.45 to 0.56, p0.001). According to the study, the overall survival probability at 5 and 10 years for patients who received chemotherapy on primary sites was about 90%, compared to only 40% and 22% for those who did not receive chemotherapy. Surgical intervention yielded similar results, but radiation therapy did not significantly improve survival rates.

**Conclusion:** Compared with men, women with SCC of the AC have a higher survival rate, and younger patients tend to live longer. The factors negatively affecting survival were increasing age, non-whites, divorced / separated / widowed, and advanced stages of cancer. A significant increase in survival was seen after surgery and chemotherapy for SCC, while radiation had little effect on survival.

## INTRODUCTION

The anal canal is defined as the caudal portion of the large intestine extending from the anorectal ring to the anal verge. Its anatomy and histology is complex and marked by the embryonic transition of endoderm to ectoderm at the dentate line.^1^ As a result of this complexity, a variety of cancers are known to originate within the anal canal. anal canal. Anal cancer constitutes less than 1% of all gastrointestinal cancers, but it is a serious public health concern.^2^ The symptoms of anal cancer can include anal pain and bleeding. Many patients with cancer of the anus or anal canal are treated with both chemotherapy and radiation, resulting in high cure rates in the early stages but increasing the risk of side effects, complications, and even death in later stages.^3–6^

Anal cancer is caused by a mutation that transforms healthy, normal cells into abnormal ones in the anal canal. As a result, cancer cells can metastasize outside of the initial tumor. Oral cancer is closely linked to sexually transmitted infections such as human papillomavirus (HPV).^7–9^ According to those studies the majority of anal cancers are HPV-positive. Risk factors for anal cancer include tobacco, alcohol, and drug use, age greater than 50 years, multiple sexual partners, and anal sex.^10^ Anal cancer can be treated by radiation therapy, chemotherapy, and surgery at an earlier stage with a higher cure rate, but with a higher risk of complications, side effects, and even death at a metastatic stage. Furthermore, cause of constipation is highly related to anal fracture which often occurs more frequently in women particularly during pregnancy and breastfeeding.^11–14^ Similarly, with increasing age, patients may suffer from complications of constipation which may ultimately increase the risk of anal cancer.^14^ Anal cancer and its tumor histology and demographic factors has been studied ^16^ and some of them introduced treatment effects.^17–19^ Some studies have been revealed on overall anal cancer mortality with disparities of cancer treatments. Therefore, this study attempts to fill the gap in our understanding of age-sex-specific mortality as well as the effects of cancer treatment on SCC in the AC to estimate overall survival probability for this disease.

## METHODS

### STUDY POPULATION

We used SEER research and 18 registries to sub-set 147 case listings released in 2020 and covering data from 2000 to 2018. It contains data on various types of cancer that have been diagnosed for approximately 28 percent of the U.S. population.^21^ The SEER Program collects population-based data on all invasive and in situ cancer diagnoses and outcomes, as well as tumor characteristics, demographics, and cancer treatments. Our focus was on the individual histologic types of SCC found in the AC, coding 8010 site recode rare tumor 12.1 in SEER. First filtering the data from site recode ICD-0-3 / WHO anus or anal canal cancer, then screening the rare tumors section 12.1 for squamous cell carcinoma with variants of anal canal. This study defined the stages based on the historical SEER stage (local, regional, or distant) and the selected treatments that were available in the dataset. Information about these data items can be found on SEER’s website (http://www.seer.cancer.gov).

### VARIABLES SELECTION

In this study, survival prediction was the primary endpoint at which to assess the efficacy of the intervention. In this study, survival time points were defined as the interval between diagnosis and death, and exposure to various cancer treatments, stages, and tumor histology, as well as demographic factors, were considered. A primary outcome variable was the listing status for patients with SCC in the AC, which was coded International Classification of Diseases for Oncology, Third Revision, [ICD-O-3] codes C210–C218.^20,21^ We used the SEER summary staging system to stage malignant, microscopically confirmed invasive tumors confined to the anal canal were defined as localized; tumors that had invaded surrounding tissues, organs, or lymph nodes were defined as regional; metastasized tumors were defined as distant. Sociodemographic variables included age, sex, race (white, and non-white; black, and other, marital status (unmarried; by state law, married, and listed separated or divorced and widowed).

### STATISTICAL ANALYSIS

Data were analyzed with R statistical software and SATA version Stata/IC 15.1 for Mac (64-bit Intel) Revision 03 Feb 2020 Copyright 1985-2017 StataCorp LLC, Single-user Stata perpetual license, Serial Number 301506386406, Licensed to Md Roungu Ahmmad, The University of Mississippi Medical Center. Pearson Chi-square test was used to determine whether there were significant differences between gender and covariates, and for continuous variables, the Wilcoxon rank-sum test was used. The observed survival rate represents the likelihood of surviving all causes of mortality over the study period. In order to estimate the overall survival rate, a univariate Kaplan-Meier method was applied, as well as a stratified Kaplan-Meier method for cancer treatment based on stages for patients with SCC. By using the log rank test, we verified whether the survival curve was statistically significant. Using unadjusted and multivariable Cox proportional hazards models, we calculated hazard ratios (HR) and 95% confidence intervals (CI) for cancer treatments and overall mortality among sexes. Finally, we inferred the overall survival probability for patients with SCC in the AC who received cancer treatments by nomograms.

## RESULTS

### PATIENTS CHARACTERISTICS

A total of 24,892 patients were included in this study (Table 1, Fig 1) where the overall median survival time was 43 months (IQR: 16 - 97) and males (38 months, IQR: 13-92) had significantly lower median survival time than females (46 months, IQR: 17-100). The age of patients ranged from 10 to 104 years where overall median age at the time of diagnosis was 60 years (IQR: 52 - 70). Among the population, whites (87%) were the largest group, but African American or American Indian men (16%) were significantly higher than white men (11%).

**Table 1:**
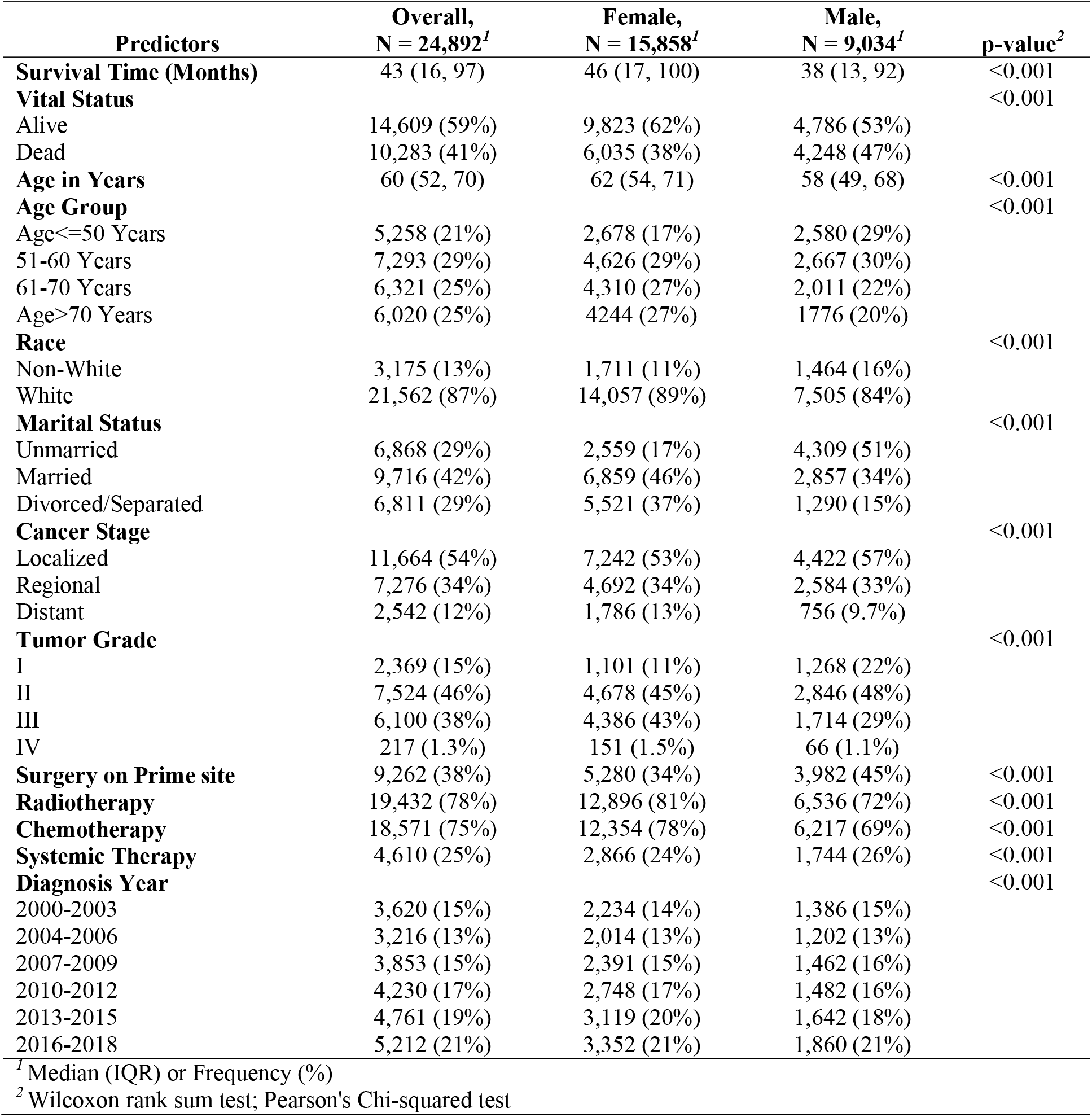
Patients Characteristics Grouped by Sex with Significance Levels.

**Figure 1:**
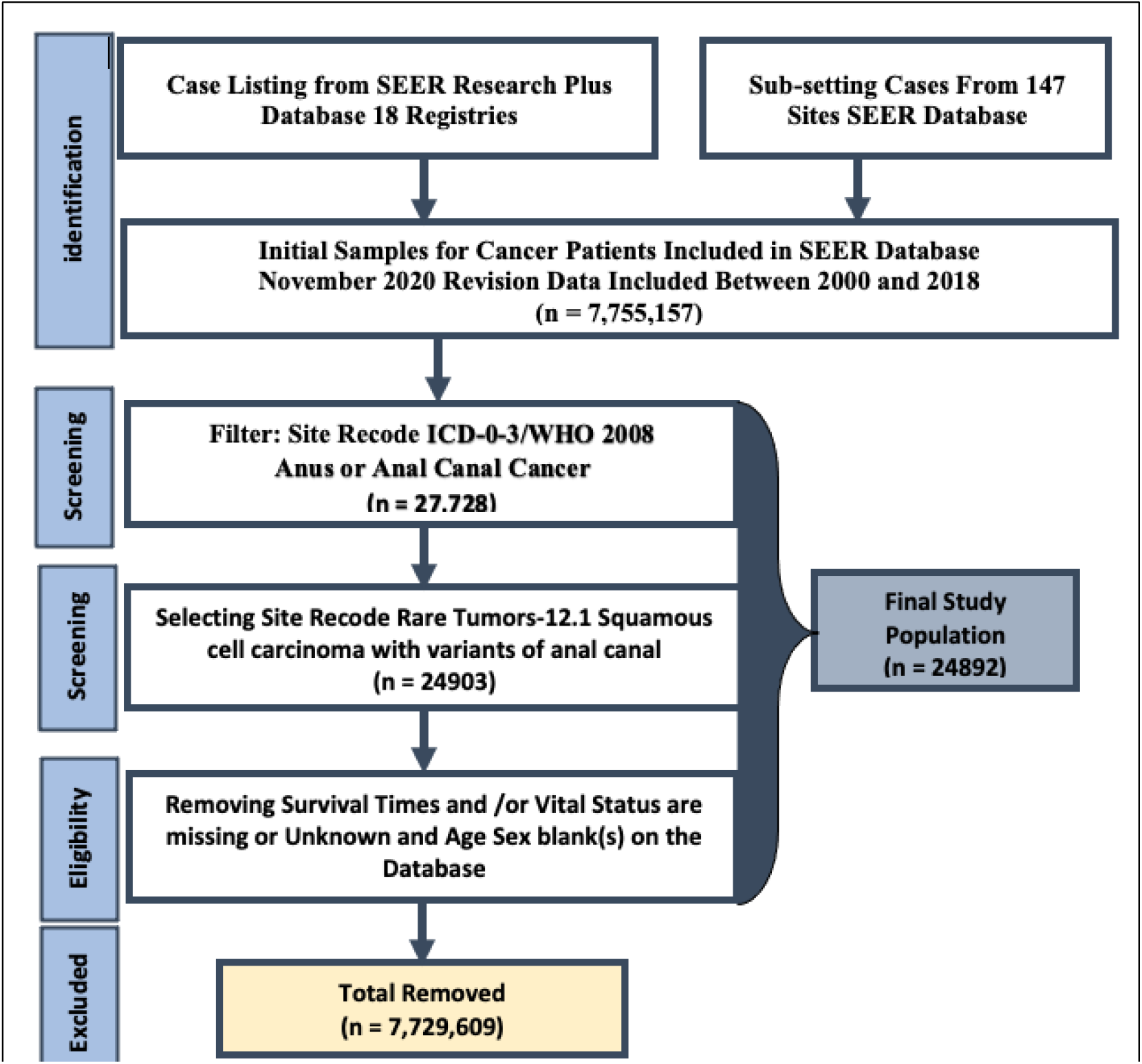
Flowchart for determination of final study populations.

Among patients with SCC, 30% were single (never married), 42% were married, and 29% were divorced, separated, or widowed, with the percentage of unmarried men being higher than that of unmarried women. However, married women and women who are separated or divorced had higher percentages of SCC in the AC. Among the SCC patients, 2,542 (12%) were diagnosed with metastatic cancer, while 7,276 (34%) and 11,664 (54%) were diagnosed with regional and localized cancer. In comparison with males, females had a significantly higher percentage of patients diagnosed at metastatic cancer stages.

The tumor grade spectrum was composed of 15% grade I tumors, 46% (n = 7524) and 38% of patients had tumor grade II or III respectively (Table 1). In the study, 70% of patients received beams or other treatments (Not shown) where as 78% of patients received radiation intervention, and 75% were given chemotherapy. A total of 9,262 (38%) patients underwent surgery on primary sites and females received a significantly higher rate of surgery than males. Among all the patients, females were more likely to undergo chemotherapy and radiation therapy than males, and only 4,610 patients (25%) had primary systemic therapy for both genders. In each of the past three years, the number of patients with the disease increased, and males and females had an equal proportion of SCC of the AC.

The impact of age, sex, and cancer treatments was assessed with Kaplan-Meier and stratified Kaplan-Meier survival curves for patients with SCC of the AC. Women have a significantly higher overall survival rate than men, and younger people tend to live longer than older people with this cancer. Statistics show that patients with distant (metastatic) stage cancer have significantly lower survival rates than patients with localized and regional stage cancer. Furthermore, white patients with SCC have significantly higher survival rates compared with non-whites. Legally married patients have a higher survival rate than unmarried or never-married patients, whereas separated, divorced, or widowed patients have a significantly lower survival rate than never-married or married patients with SCC in the AC (Fig.2). More specifically, patients with metastatic stages have a six-year survival rate of 25%, regional stage patients 55%, and local stage patients 75% (p < 0.001, Fig. 2, P4). Cancer patients with SCC in the AC benefited from radiotherapy, chemotherapy, and surgery on the primary site (Fig. 2: P7-P9). Patients with metastatic cancer stages who were treated with radiation had a 2-year overall survival rate of 55% compared to 20% in those who did not receive radiation intervention (Fig. 2, P7). Additionally, the 2-year overall survival rate for patients with distant stage disease who received chemotherapy on the primary site was above 75% as compared to 20% for those who did not receive chemotherapy on the primary site (p < 0.001, Fig. 2, P8-9). In short, chemotherapy on the primary site improved 55% of survival rates for patients with SCC in the AC, whereas radiation intervention improved only 35%, and surgery did not significantly improve survival rates. Therefore, chemotherapy and radiotherapy significantly improve survival for the patient with SCC, but surgery on the primary site did not significantly improve survival.

**Figure 2:**
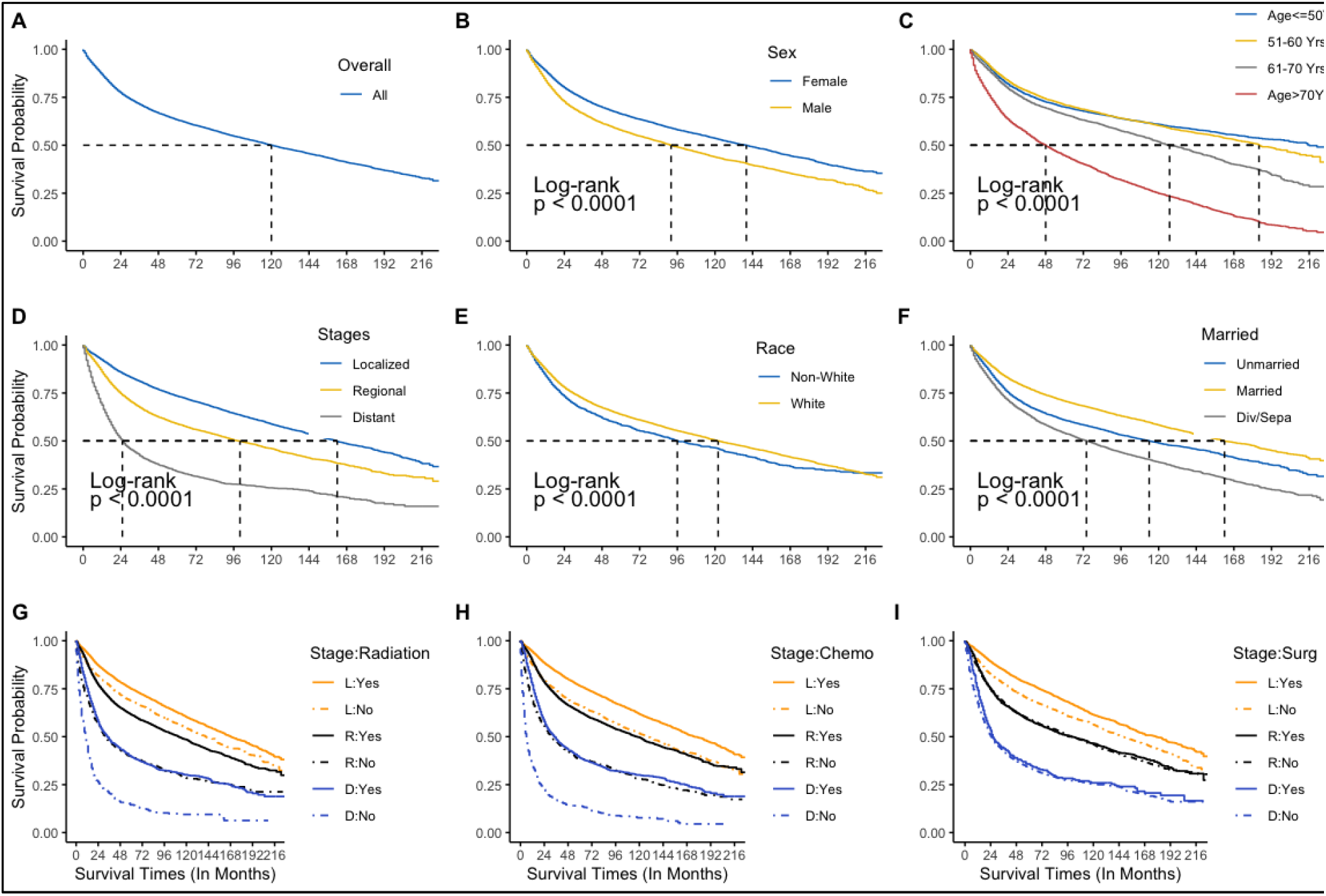
Univariate survival probability for the patients with SCC at the AC. A) Overall Survival probability B) survival probability among Sex C) among Age group D) among cancer stage E) among race F) marital status G) cancer stage and radiation H) stage and Chemo and I) stage and surgery.

The factors negatively affecting survival on multivariate analysis were increasing age, male sex, non-whites, advanced cancer stages, and tumor grade **(Fig. 3)**. Compared to the untreated population, radiotherapy, chemotherapy, and surgery significantly decreased the risk of mortality, whereas systemic therapy had the opposite effect on multivariate analysis. Married patients had significantly lower mortality rate compare with unmarried or never-married patients (HR: 0.76, 95% CI: 70-83, p<0.001) but divorced, separated, or widowed patients had 16% higher risk of death compare with unmarried. Additionally, married patients with SCC have a significantly lower risk of death when compared with divorced, separated, or widowed patients. In metastatic stages, patients had a 3.34-fold higher hazard of death compared to patients in earlier stages (HR 3.27, 95% CI: 3.05 – 3.64, p<0.001). For the those with intermediate (regional) stage disease, patients had 65% higher risk of death compared to localized stages (HR: 1.64, 95% CI: 1.52-1.76, p<0.001). In adjusting for demographics and other predictors, the patients who underwent surgery on the primary site had a 49% (HR:0.51, 95% CI: 0.46-0.56, p<0.001) lower risk of death compared with those who did not. Additionally, the patients who received chemotherapy at the prime site had a 55% lower risk of death (HR:0.51, 95% CI: 0.40-0.50, p<0.001) than those who did not. Finally, those who received radiation therapy at the primary site had a 26% lower hazard of death (HR:0.74, 95% CI: 0.67-0.83, p<0.001) than those who did not receive radiation intervention. Therefore, surgical intervention on the primary site, radiation intervention, and chemotherapy significantly reduced the risk of mortality, but primary systemic therapy provided contradictory results.

**Figure 3:**
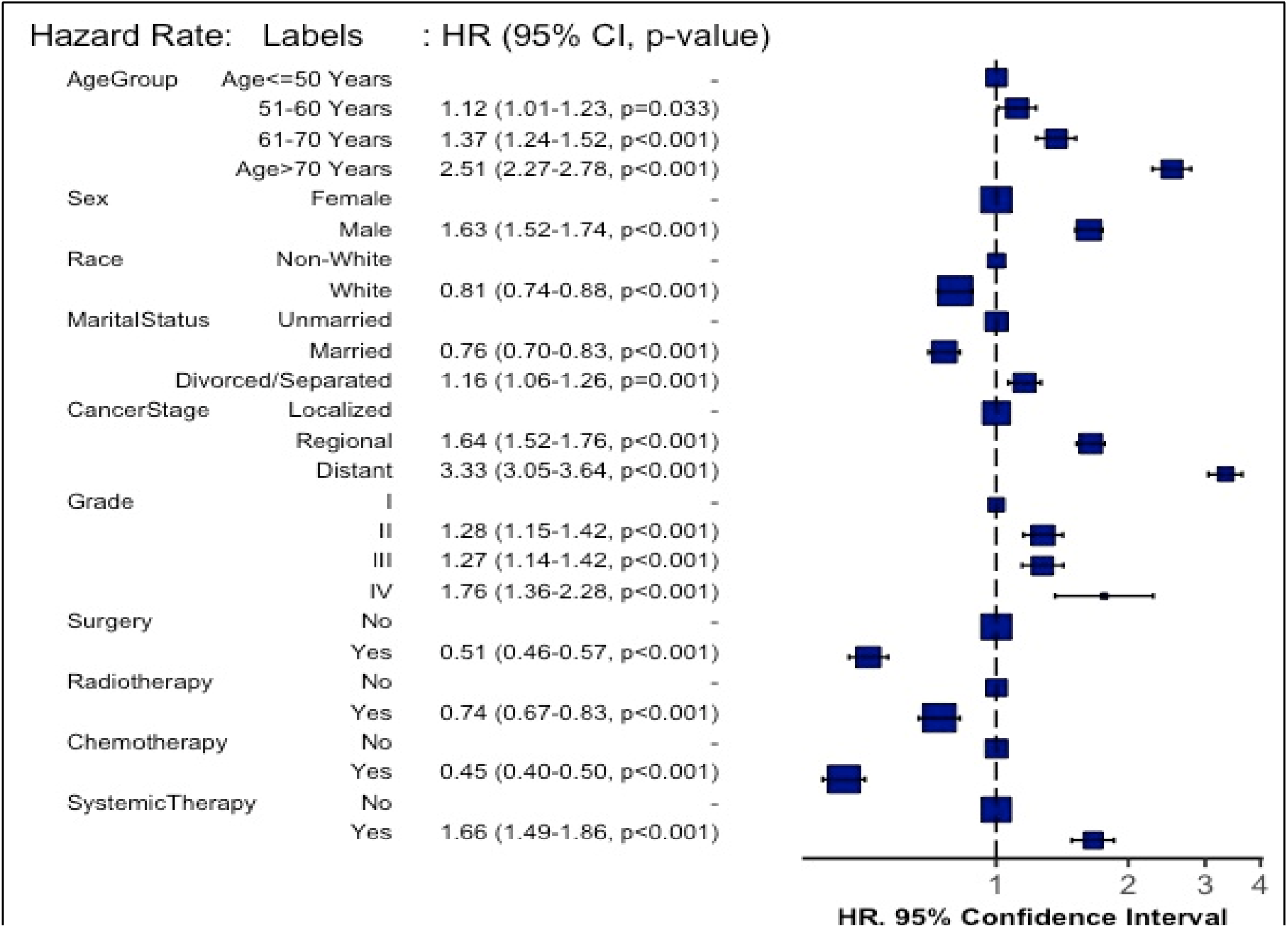
Overall hazard rate for the patient with SCC at the AC and corresponding forest plots.

### WHY AGE-SEX SPECIFIC ANALYSIS NEEDED

On average, the number of patients with SCCs of the AC has increased significantly over time, and with the consideration of genders, the female population has a higher percentage (n = 15,858) than the male population (n = 9034) in this study. However, the death rate for males (47%, n = 4,248) among the study population was higher than for females (38%, n = 6,035). In our study population, over 21% (n = 5,258) were under the age of 50 with the majority being in the 51-60 years age group. Some literature suggests that sex is an important factor contributing to constipation that may lead to anal fractures.^22–24^. Women are susceptible to constipation during especially during pregnancy or breastfeeding,.^22,23^ However, males are not affected by this type of biological process. Several studies have shown that males lead more sedentary lives than females.^25^ According to NCI reports, sex is an important factor in anal cancer due to some biochemical factors. In general, the elderly have more frequent gastrointestinal disorders and may be more prone to issues with constipation than younger people.^26–28^ Age is another significant factor in anal cancer, according to NCI reports. According to a Medscape report, females have a 3 times higher rate of constipation than males in the United States, but other studies showed females have overall higher survival than males. ^27^ The incidence of anal cancer is associated with a number of factors, including constipation, anal sex, age, and chronic diseases.^12,29,30^ Several studies revealed that the overall 5-year incident rate for female is higher compare with males, but different outcomes have been found between 15-49 years old.^1,31^. According to this study, the overall proportion of females with SCC in AC was higher than the proportion of males, and females aged 15-49 years had a higher proportion than their male counterparts. Furthermore, the Cox model indicated that overall male patients tend to have a higher rate of mortality than female patients do. Women are more likely to have behavioral and reproductive activities while they are younger than 49 years of age. Therefore, in view of this, it is necessary to investigate further into the mortality amongst different sexes and age groups.

For both adjusted and unadjusted predictors, the results for stratified Cox proportional hazards models are shown in table 3 and figure 3. For a single year increase in age, mortality increased 5% for female patients (HR: 1.05, 95% confidence interval: 1.04-1.05, p<0.001 for unadjusted model) and 4% increased by adjusting predictors, on the other hand for male has only 3% and 2% respectively. For grouped ages, women 51-60 years have 18% (HR: 1.18, 95% CI: 1.02-1.37, p=0.030) higher hazard than those below 51 years, while males have only 7% higher hazard. Moreover, females are more likely to die as they age than males.

**Table 3:** Hazard rate stratified by Age group with adjusted and unadjusted predictors for the patients with SCC at the AC.

| Predictors | Age greater than 49 Years |  | Age less than 50 Years |  |
| --- | --- | --- | --- | --- |
|  | Unadj. HR (95% CI, p) | Adj. HR (95% CI, p) | Unadj. HR (95% CI, p) | Adj. HR (95% CI, p) |
| Sex: Female | Ref | Ref | Ref | Ref |
| Male | 1.35 (1.29-1.41, p<0.001) | 1.60 (1.48-1.72, p<0.001) | 1.72 (1.56-1.90, p<0.001) | 1.90 (1.55-2.33, p<0.001) |
| Race: Non-White | Ref | Ref | Ref | Ref |
| White | 0.90 (0.84-0.96, p=0.002) | 0.94 (0.84-1.04, p=0.218) | 0.61 (0.55-0.68, p<0.001) | 0.58 (0.48-0.71, p<0.001) |
| Marital Status: Unmarried | Ref | Ref | Ref | Ref |
| Married | 0.74 (0.70-0.79, p<0.001) | 0.85 (0.78-0.93, p<0.001) | 0.52 (0.46-0.59, p<0.001) | 0.73 (0.58-0.92, p=0.009) |
| Divorced/Separated | 1.27 (1.21-1.35, p<0.001) | 1.46 (1.34-1.60, p<0.001) | 0.85 (0.74-0.98, p=0.030) | 1.02 (0.79-1.33, p=0.869) |
| Cancer Stages: Localized | Ref | Ref | Ref | Ref |
| Regional | 1.50 (1.43-1.58, p<0.001) | 1.56 (1.44-1.69, p<0.001) | 1.78 (1.59-1.99, p<0.001) | 1.99 (1.61-2.45, p<0.001) |
| Distant | 3.06 (2.88-3.26, p<0.001) | 3.11 (2.82-3.42, p<0.001) | 3.68 (3.20-4.24, p<0.001) | 4.88 (3.78-6.31, p<0.001) |
| Tumor Grades: I | Ref | Ref | Ref | Ref |
| II | 1.19 (1.10-1.29, p<0.001) | 1.28 (1.14-1.43, p<0.001) | 1.18 (1.00-1.38, p=0.044) | 1.40 (1.09-1.81, p=0.009) |
| III | 1.21 (1.11-1.31, p<0.001) | 1.30 (1.15-1.46, p<0.001) | 1.30 (1.09-1.54, p=0.003) | 1.69 (1.28-2.24, p<0.001) |
| IV | 1.48 (1.20-1.83, p<0.001) | 2.04 (1.56-2.68, p<0.001) | 0.98 (0.57-1.68, p=0.936) | 1.14 (0.49-2.64, p=0.756) |
| Surgery | 0.74 (0.71-0.78, p<0.001) | 0.49 (0.44-0.55, p<0.001) | 0.67 (0.61-0.74, p<0.001) | 0.61 (0.45-0.83, p=0.002) |
| Radiotherapy | 0.64 (0.61-0.67, p<0.001) | 0.73 (0.65-0.81, p<0.001) | 0.92 (0.82-1.03, p=0.144) | 0.88 (0.64-1.21, p=0.425) |
| Chemotherapy | 0.59 (0.57-0.62, p<0.001) | 0.40 (0.35-0.45, p<0.001) | 0.93 (0.83-1.04, p=0.192) | 0.43 (0.30-0.61, p<0.001) |
| Systemic Therapy | 0.70 (0.66-0.75, p<0.001) | 1.68 (1.49-1.89, p<0.001) | 0.90 (0.78-1.04, p=0.152) | 1.45 (1.06-1.98, p=0.021) |

The mortality rate of female patients with SCC who underwent surgery on the primary site was reduced by 42% (HR 0.58, 95% CI: 0.51-0.66, p<0.001) compared to those who did not receive surgery by adjusting other predictors (Tab 3), and that of males was reduced by 57% (HR: 0.43 (0.37-0.51, p<0.001). Similarly, the mortality rate of female patients with SCC who underwent chemotherapy was reduced by 51% (HR 0.49, 95% CI: 0.42-0.56, p<0.001) compared to those who did not receive chemotherapy by adjusting other predictors (Tab 2) and that of males was reduced by 60% (HR: 0.40, 95% CI: 0.34-0.48, p<0.001). Patients at metastatic stage (distant) have a higher risk than those at localized or regional stages both for females and males (Fig. 3, Tab. 3). Comparing the white population to non-whites, among the sexes, the white population has lower mortality than the non-white population. Additionally, marital status proved an important indicator for the development of anal canal cancer. The mortality rate of female patients with SCC who were married was 24% lower (HR 0.76, 0.67-0.86, p<0.001) than those who were unmarried (Tab 2). Contrastingly, divorced/separated female had 18 % higher hazard of death compare with single women.

Significant findings have been found when grouped by age in 10-49 years and above 49 years old patients. The female patients in the 10-49 year old age group had 41% lower risk of mortality compared with those patients greater than 49 years (HR: 0.59, 95% CI: 0.51-0.68, p<0.001) by adjusting cancer treatment and other predictors. Males had 31% and 23% respectively lower risk of death for unadjusted and adjusted models compare with older patients (Tab App 1). Thus, young women with SCC of the AC are at a higher risk of mortality than men. Furthermore, married women’s hazard rate was higher than married men’s hazard rate compared with those who were unmarried or never-married, but divorced and separated men tended to have a lower hazard rate in those groups than single or never-married men [Fig 3]. In addition, males have a higher mortality rate during metastatic stages as compared with females during localized and regional stages. Surgical intervention on the primary site resulted in a lower mortality rate for males than females, but radiation intervention provided opposite outcomes among sexes [Fig 3]. The efficacy of chemotherapy for males and females was approximately the same, but the effectiveness of primary systemic therapy was contradictory.

Fig 5 represented the nomogram associated with the multivariate Cox proportional hazard model predicting the overall survival probability for patients with SCC in the AC by adjusting predictors. Assigning a point value to each variable is accomplished by first locating the patient’s characteristics row and then drawing a vertical line straight up to the points row. By drawing a vertical line directly up to the point row in Fig 4, we obtain a score of 35 points for the patient age above 49 years. Repeat this process for each row, accumulating points for each variable. The overall survival probability is determined by adding up all the points and drawing a vertical line from the total points column. For example, if each variable contributes 140 points, then the overall survival probability at the end of 5 years would be 70%, and 10 years would be 55%. Also, patients who had surgery on prime sites survived approximately 90% and 80% for 5 and 10 years, whereas those without surgery only lived 25% and 10%. Similarly, radiation intervention did not improve survival a lot but chemotherapy on the SCC on the AC improve survival significantly. The primary systemic therapy for this cancer, however, did not seem to be effective in improving survival.

**Figure 4:**
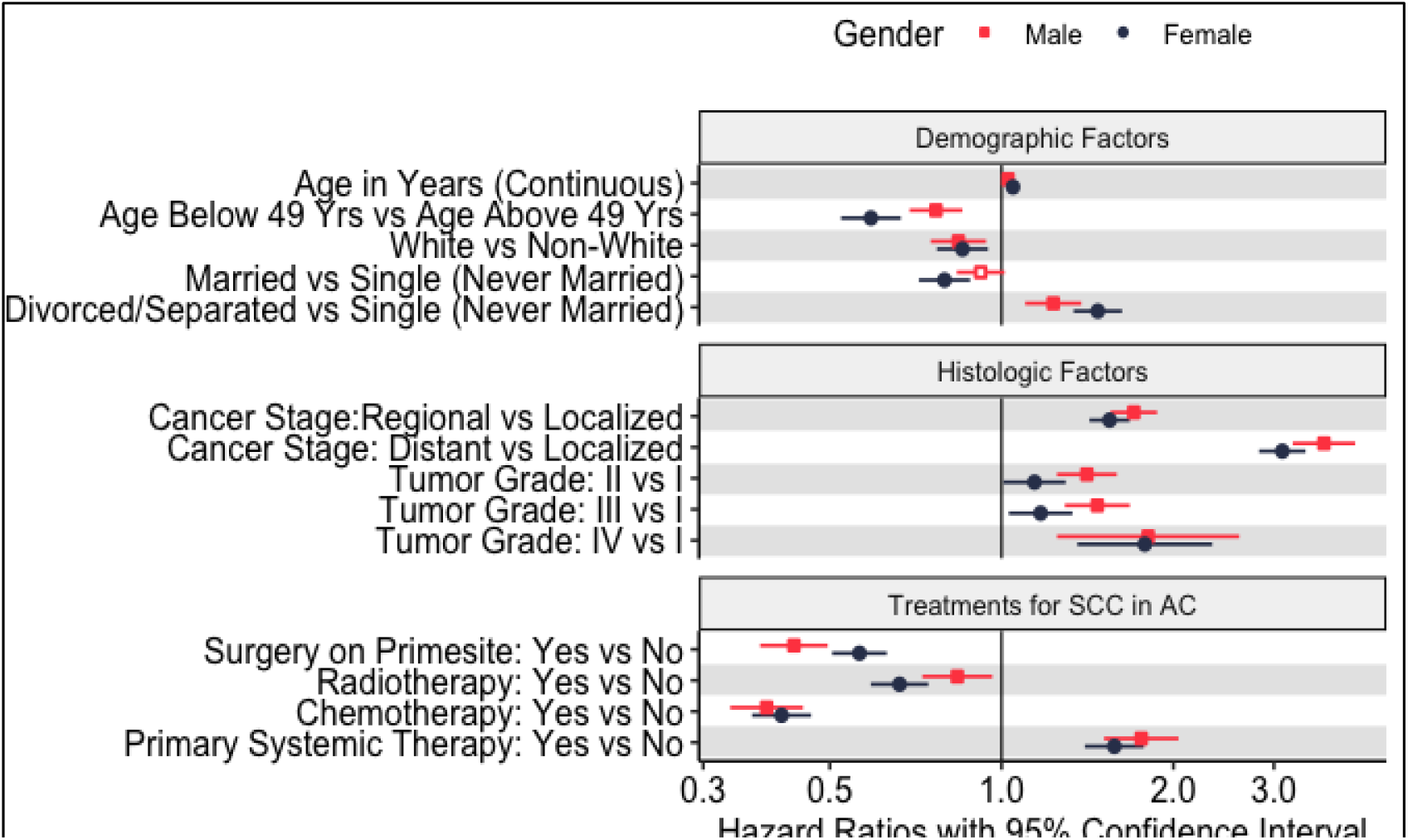
Adjusted hazard rate corresponding hazard forest plot grouped by sex for the patients with SCC at the AC.

**Figure 5:**
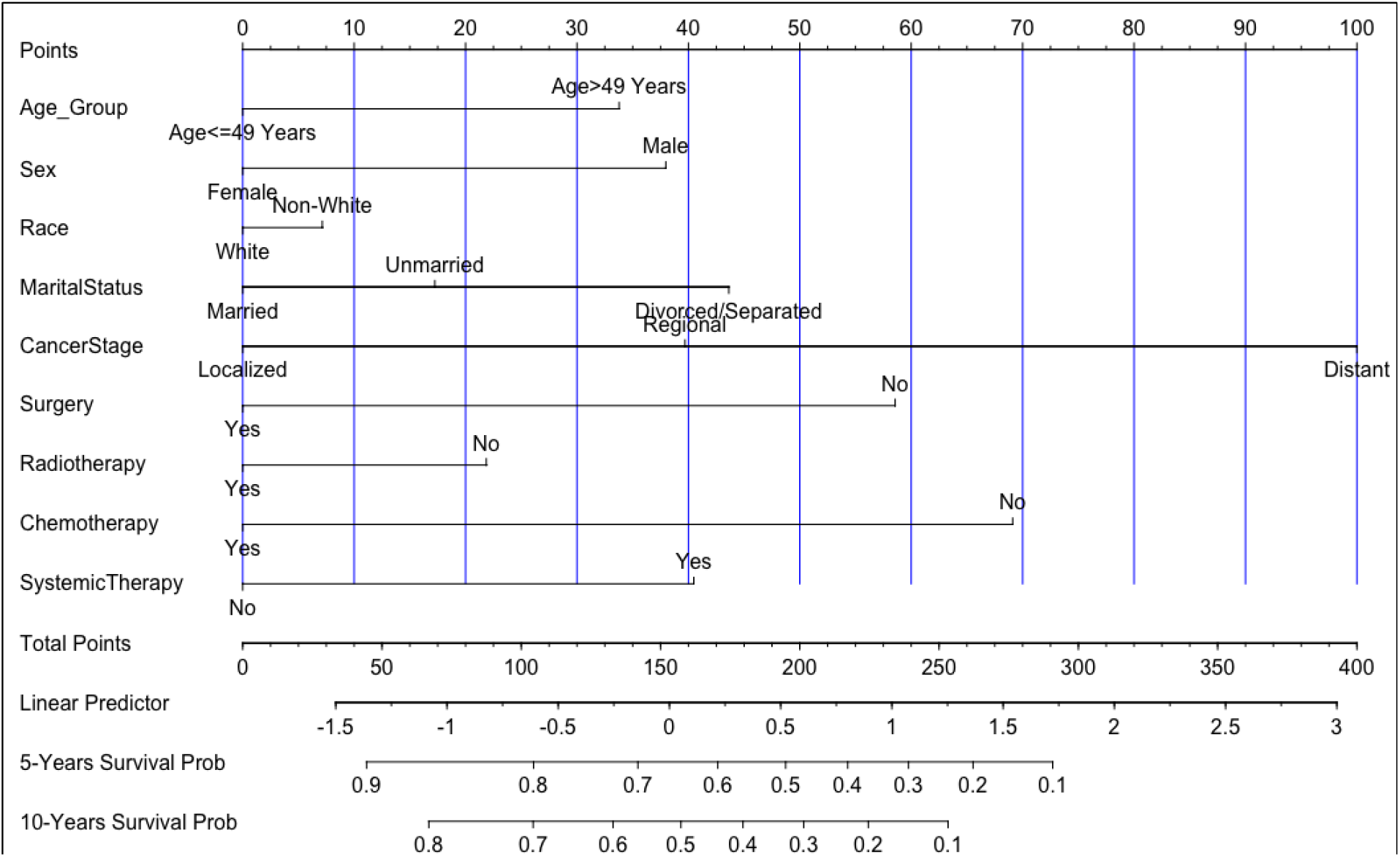
Overall survival nomogram. In this nomogram, we provide a method for calculating the five- and ten-year probability of survival based on the combination of covariates of a patient. Draw a straight line down the 5-year and 10-year axes to calculate the probability.

## DISCUSSION

This study investigated the impact of cancer treatment on patients who were diagnosed with SCC of the AC as a part of a large population-based cohort study based on SEER data. Our study differs from others due to the study populations, follow-up times, and the methodology used for modeling. To predict survival and morality among age groups and sexes, we applied Kaplan-Meier methods and a stratified Cox model for patients with SCC in the AC. According to our knowledge, this is the first study that investigates the age-sex specific mortality predictions. In our study, we identified 24,892 patients with SCC in the AC with a median survival time of 43 months (IQR: 16-97), where female (46, IQR: 17-100 months) patients had higher median survival time than male (38, IQR: 13-92 months) patients. The average age of patients diagnosed with SCC in the AC was 60 years old where 21% were aged below 50 years and most of them were white married and received cancer treatments. According to this study, age, gender, race, marital status, cancer treatments, and stages had an important effect on mortality for patients with SCC of the AC.

Many studies have demonstrated that the marital status of patients with anal canal cancer has a strong association with mortality.^36,37^ This study is an extension of those studies, examining squamous cell carcinoma of the annal canal, and identifying that married patients have a higher survival rate than single or never-married, and separated or divorced patients respectively. Moreover, married patients were more likely to be diagnosed with this disease than unmarried, divorced, or separated patients. On average, half of patients with an initial diagnosis were in localized stages (54%), whereas 12% of the patients were in metastatic stages. In previous studies, patients with metastatic stages of squamous cell carcinoma had a lower survival rate than those with earlier stages ^38,39^ and our study has come to a similar conclusion. By stratifying Kaplan-Meier methods, we determined that chemotherapy improved patient survival significantly in the metastatic stage in comparison to those who did not chemotherapy at the primary site. Furthermore, radiation intervention reduced mortality, but surgery on the primary site did not significantly improve survival. According to NCI reports, anatomical canal cancer affects men and women differently based on their age and sexuality **[ref]**. According to this study, men have a higher mortality risk than females [HR: 1.62, 95% CI: 1.52-1.74, p<0.001] which is consistent with previous studies. Several studies demonstrated radiation therapy, chemotherapy and surgical intervention improved overall survival for patients with rectum and colon cancer.^40–45^ The Cox proportional hazard model was fitted to the entire study, and it was found that chemotherapy, radiation intervention, and surgery at the primary site were significantly associated with mortality; however, primary systemic therapy failed to provide positive results [figure 4].

In this study, stratified Cox proportional hazard models were fitted to predict death rates among patients who underwent surgery, radiation therapy, chemotherapy, and systemic therapy. According to our study, the mortality rate for patients with SCC of the AC increases with age among sexes. Among those between 15-49 and older than 49, the mortality rate for females is lower than for males. The chemotherapy, surgery, and radiation intervention at the primary site resulted in a lower mortality rate for patients with SCC of the AC, which is consistent with other studies.^46–48^ While radiotherapy and systemic therapy may benefit patients with this illness, some studies suggest they may not be able to improve survival when in advanced stages (SCC).^40^ According to this study, men tended to die less from chemotherapy and surgery on the primary site than women, while the opposite was true for radiation therapy interventions. Studies have employed nomograms in the prediction of survival for breast cancer, prostate cancer, melanoma, thyroid cancer, and head and neck squamous cell carcinoma, based on different study phases.^32–35,49–52^ This study is the first one to employ a nomogram as an instrument to predict the survival of patients with SCC in the AC. Furthermore, our model and nomogram were easy to understand by clinicians, which is another strength of the study. Our study showed that patients who received chemotherapy at the primary sites survived longer than those who did not receive chemotherapy. Chemotherapy is more effective than surgery and radiation on patients with SCC in the AC. The use of systemic therapy provided opposite results owing to a lack of power or small sample size in this study, which may have contributed to contradictory conclusions.

Despite using a large dataset from SEER, we found some limitations in our study. In the absence of public access to SEER, some clinicopathological factors (such as surgical margins, perineural invasion, solid tumors, and p53 positivity) could not be examined. Furthermore, the SEER public-use dataset does not contain any data on the histological subtypes of SCC in the AC, despite the importance of histological details in predicting survival. Additionally, SEER does not contain information about cancer recurrences. There was no information on radiotherapy technique (total dose, fraction size, beam energy), so the results were unable to demonstrate whether survival is affected by these factors. Furthermore, although this study did not include some confounding factors (such as tumor depth, margin, and performance status), it adjusted for all patient and tumor characteristics. Radiation might be justified for certain low-grade sarcomas if certain risk factors (i.e., incomplete resection, positive margins) were not found in SEER. Another drawback of this study is the relatively short follow-up period, as only a few patients were available for the duration of the study. A longer follow-up may improve the accuracy of the model, especially in patients with SCC in the AC. Though the model and nomogram performed well, external validation with other cohorts is still necessary to confirm the efficacy of the model.

## CONCLUSION

The study was one of the most comprehensive population-based cancer surveys to be conducted in the United States. Women and white patients tend to experience higher survival rates than their counterparts. Unmarried, divorced, and separated patients have a lower survival rate than married patients. Patients with SCC had a better survival outcome after receiving surgery, radiation therapy, and chemotherapy, although primary systemic therapy had conflicting results. Radiation intervention did not have a big impact on survival in this study, but surgery, as well as chemotherapy used on the SCC, improved survival by a substantial margin.

## Data Availability

All data produced in the present study are available upon reasonable request to the authors.
Raw all data produced are available online at https://seer.cancer.gov/about/

## ACKNOWLEDGEMENT

The authors would like to thank the NCI for open access to their SEER database. The opinions or views expressed in this paper are those of the authors and do not represent the opinions or recommendations of the NCI.

---

**Table App 1:**
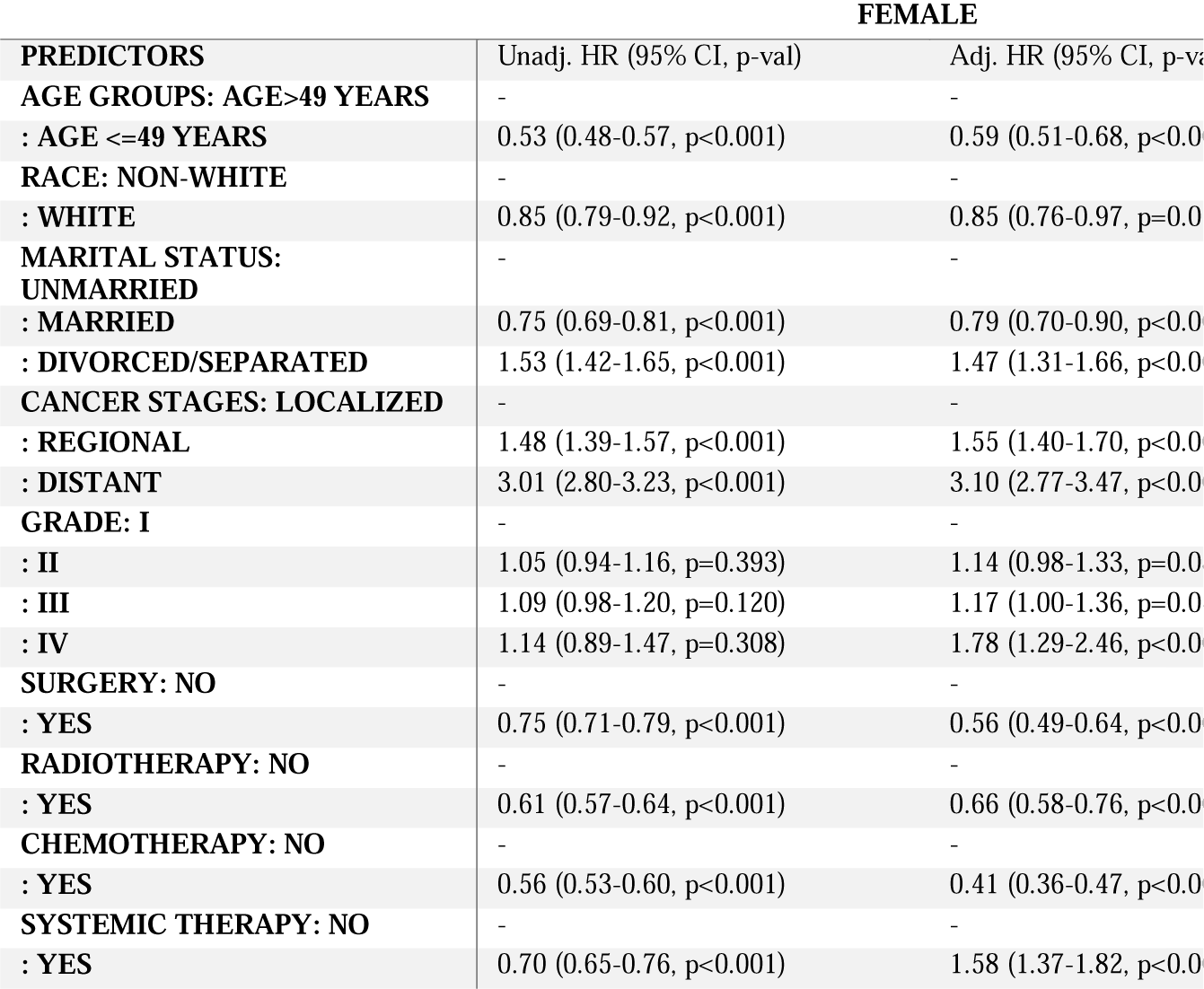
Stratified Cox model with Sex-Specified outcomes.

